# Multiplex Pan-Filovirus Assay Performance and Reproducibility Across Varied Geographical and Resource Settings

**DOI:** 10.64898/2026.05.07.26352689

**Authors:** Olivia A. Smith, Sydney Merritt, Jean Paul Kompany, Nicole A. Hoff, Teri Ann S. Wong, Varney Kamara, Merly Tambu, Megan Halbrook, Jason Kindrachuk, Angelica L. Barrall, Kamy Musene, Skylar A. Martin, John Berestecky, Robert Orr, Todd Myers, Tracy MacGill, Jean-Jacques Muyembe-Tamfum, Didine Kaba, Placide Mbala-Kingebeni, Anne W. Rimoin, Axel T. Lehrer

## Abstract

Multiplex bead-based immunoassays (MIAs) are promising tools for simultaneously detecting humoral immunity to multiple targets, potentially playing a crucial role in serosurveillance and vaccine response assessments. However, evaluation of assay performance is paramount prior to widespread use. This study presents a performance evaluation of a pan-filovirus MIA through characterization of the analytical range for the EBOV glycoprotein (GP) target and assessments of assay precision and antigen discrimination. The precision of the MIA was evaluated by comparing the detection of anti-filovirus antibodies at two independent laboratory sites: the University of Hawai’i, Honolulu (UH), and the Institut National de Recherche Biomédicale (INRB) in Kinshasa, Democratic Republic of the Congo (DRC). Forty-six samples from Yambuku, DRC, including Ebola virus Disease (EVD) survivors and close contacts, were tested at both sites. Additionally, 858 samples were tested in DRC before and after vaccination with a prophylactic EVD vaccine, ERVEBO. Results demonstrated low variability between laboratories, with intra-assay and inter-laboratory coefficients of variation below predefined thresholds for all filovirus targets included in the multiplex panel. Analyte correlations between sites were high (r^2^=0.86-0.92). Longitudinal analysis detected increased EBOV GP reactivity following vaccination, while reactivity to non-vaccine filovirus antigens remained stable, consistent with minimal cross-reactivity in a vaccinated cohort. These findings suggest that this pan-filovirus MIA produces reproducible results across distinct laboratory settings and may serve as a useful tool for comparative serologic investigations, serosurveillance, and evaluation of EBOV vaccine-associated antibody responses.

**IMPORTANCE STATEMENT:** This study investigates the functionality and intra-laboratory consistency of a novel multiplex bead-based immunoassay with pan-filovirus targets. As part of the evaluation process, the feasibility of using the assay in resource-limited settings was demonstrated in the Democratic Republic of the Congo. This assay holds significant promise as a tool for detecting filovirus-specific antibody responses. By leveraging its multiplex capabilities, it may be used for widespread serosurveillance of high-consequence pathogens, including the pan-filovirus antigens such as Ebolavirus and Marburg virus already incorporated in the assay, as well as other targets of interest.

## INTRODUCTION

Given the disproportionate impacts of both communicable and non-communicable diseases in low-resource settings, there is an increasing need for expanded serological assessments, diagnostic testing, and healthcare access. Beyond access, assurance of assay reproducibility and performance across disparate global settings is of critical importance to seroepidemiology and vaccine deployment investigations in low-and middle-income countries (LMICs). Quantitative assay evaluations as a part of validation studies typically include assessments of assay precision, specificity, linearity, and overall integrity (1–3). These parameters include determination of assay-and antigen-specific reactivity cutoffs and assessment of intra-and inter-assay variability.

For longitudinal investigations, evaluation of assay performance increases the complexity of the validation process. Incorporation of diverse sample types, including pre-and post-vaccination, or specimens from suspected or confirmed pathogen exposures, shifts performance assessment from static validation to a dynamic evaluation that accounts for the longitudinal biological variability inherent in field-based research. Such settings often include rapid vaccine deployment, cold-chain and long-term storage challenges, and extended immune profiling among diverse populations. In this context, serological investigations have become a central component of global health research and response activities for both emerging pathogens and vaccine-preventable diseases, including evaluation of vaccine-induced immune responses and assessment of antibody durability over time.

Widespread serological testing in LMICs can improve in-country detection of known pathogens and enhance understanding of infectious disease landscapes in regions that historically have relied heavily on clinical diagnoses and symptom presentation (4). Conducting testing in-country may also enhance research autonomy through decentralization of laboratory testing and local capacity building. Further, in-country testing can reduce the risks to sample integrity associated with transportation and prolonged storage. Multiplex bead-based immunoassays (MIAs) have gained substantial interest as tools for investigating humoral immunity and antigen-specific exposure (5). However, despite their growing use, systematic evaluations of MIA performance across geographically and resource-diverse settings remain limited. Thus, performance evaluations of these assays in resource-limited settings would support broader implementation and expansion of serological testing capacity globally.

The *Filoviridae* family includes several high-consequence virus species associated with high morbidity and mortality in humans, most notably *Orthoebolavirus zairense* (EBOV) and *Orthomarburgvirus marburgense* (MARV) (6). EBOV, the causative agent of Ebola virus disease (EVD), was first identified in 1976 from the Yambuku region of the Democratic Republic of the Congo (DRC) and has driven unpredictable outbreaks across Central and West Africa over the following decades (7, 8). Vaccine design and development strategies, including the VSV-vectored recombinant EBOV-GP vaccine, ERVEBO, have targeted the viral glycoprotein (GP) and elicit a sustained humoral immune response including the development of anti-GP immunoglobulin G (IgG) antibodies (9–11). In addition to GP, the nucleoprotein (NP) and viral matrix protein 40 (VP40), are also primary targets of antibodies, (12), and may provide evidence for prior virus exposure (13).

While MIAs offer high-throughput flexibility, we specifically sought to validate a multiplexed panel targeting filovirus-specific IgG. This approach enables the differentiation of vaccine-induced immunity (GP-focused) from signatures of natural infection (incorporating NP and VP40) A key advantage of the multiplex approach is its ability to simultaneously detect responses to multiple antigens, enabling characterization of antigen-specific reactivity patterns among survivors of natural EVD infection and individuals vaccinated against EBOV. Using MAGPIX technology, we present a performance evaluation of a multiplex filovirus MIA based on the detection of filovirus-specific IgG responses in a diverse population of EBOV-exposed and ERVEBO-vaccinated individuals under real-life conditions.

## MATERIALS AND METHODS

### Parameters of Assay Evaluation

#### Assay Precision and Reproducibility

Assay precision was evaluated by assessing intra-assay and inter-laboratory variability using human serum samples from Cohort 1. A total of 46 samples were included, comprising three sera from confirmed EVD survivors and 43 sera from individuals without documented EVD history. Each sample was tested in duplicate at two independent laboratory sites: the University of Hawai’i (UH) and the Institut National de Recherche Biomedicale (INRB) in Kinshasa, DRC.

Intra-assay variability was calculated as the coefficient of variation (CV) between duplicate measurements within the same laboratory. Inter-laboratory variability was assessed by comparing mean analyte-specific median fluorescence intensity (MFI) values obtained at UH and INRB for each sample. An acceptance criterion of CV <= 15% was used for both intra-assay and inter-laboratory precision assessments (14–16).

#### Linearity and Analytic Range

Analytic linearity and range were evaluated using a non-human primate (NHP) derived EBOV GP-specific IgG standard. Serial dilutions of the purified EBOV GP-binding IgG standard were used to generate a sigmoidal dose-response curve, from which the linear portion of the curve was identified (r^2^ > 0.99). The lower and upper limits of quantification (LLOQ and ULOQ) were defined based on the lowest and highest concentrations within this linear range. Linearity and quantitative performance were assessed only for the EBOV GP analyte using the NHP standard. Quantitative interpolation was not performed for human serum samples, and no standards were available to evaluate linearity or analytical range for non-EBOV filovirus antigens included in the multiplex panel.

#### Specificity and Cross-Reactivity Assessment

Specificity of antibody detection was evaluated using sera from Cohort 2, which included individuals vaccinated with recombinant vesicular stomatitis virus-Zaire Ebola virus glycoprotein (rVSV-ZEBOV-GP) vaccine, ERVEBO. These samples were collected longitudinally before vaccination and at multiple time points post-vaccination. Specificity was assessed by comparing antibody reactivity to EBOV GP with reactivity to other filovirus antigens included in the multiplex panel (MARV GP, SUDV GP, and BDBV GP). As ERVEBO is expected to elicit antibodies specific to EBOV GP only, sustained increases in EBOV GP reactivity in the absence of corresponding increases to non-EBOV filovirus antigens were interpreted as evidence of assay specificity and minimal cross-reactivity. Sera from individuals with reported histories of infection with MARV, SUDV, or BDBV, as well as sera from individuals with other acute viral infections, were not included in specificity or cross-reactivity analyses.

#### Serum Samples

Human serum samples used for assay evaluation were derived from two cohorts collected in the DRC. **Cohort 1** consisted of 46 serum samples collected in 2016 from individuals in Yambuku. DRC, including three samples from confirmed EVD survivors from the 1976 Yambuku outbreak and 43 community members without documented EVD history at the time of sampling. These samples were selected from a larger parent cohort and were used exclusively for precision and inter-laboratory reproducibility analyses. All 46 samples were tested in duplicate at two independent laboratory sites. Samples were de-identified prior to analysis.

**Cohort 2** included 858 serum samples collected in 2018 from two EVD-affected regions in DRC: Mbandaka, Equateur Province (n = 395) and Beni, North Kivu Province (n = 463). Samples were obtained both before and after vaccination with the ERVEBO, as well as from close contacts of confirmed or suspected EVD cases and healthcare workers. These samples were used to evaluate longitudinal antibody responses and assay specificity and were analyzed only at the INRB.

All samples were collected under protocols approved by the University of California, Los Angeles Institutional Review Board (IRB Nos. IRB15-000333 and IRB16-001346) and the Kinshasa School of Public Health (KSPH) IRB (IRB Nos. ESPCE0382015 and ESPCE0222017). Serum samples were stored frozen at-80 °C and thawed once before testing. Human sera from known survivors of BDBV, SUDV, or MARV were not included in the analyses.

#### Multiplex Assay

Magnetic MagPlex microspheres (Luminex, Austin, TX), internally dyed, were covalently coupled with purified-proteins from multiple filoviruses including EBOV GP, EBOV viral protein 40 (VP40), EBOV nucleoprotein (NP), BDBV (*Orthoebolavirus bundibugyoense*) GP, SUDV (*Orthoebolavirus sudanense*) GP, MARV GP, and MARV VP40 expressed in *Drosophila* S2 cells and purified at UH. Bovine serum albumin (BSA)-coupled beads were included as a negative control to assess nonspecific serum binding, as previously described (5, 17–19).

Beads were coupled at UH using standard carbodiimide chemistry, combined into multiplex panels, and shipped to the INRB. Both laboratories used beads derived from the same coupling lots, thus lot-to-lot variability was not evaluated. Beads were stored at 4 °C in the dark, and while beads were not formally requalified post-shipment, the high concordance between UH and INRB sites (figure 1) served as a functional validation of bead stability during transport.

**Figure 1:**
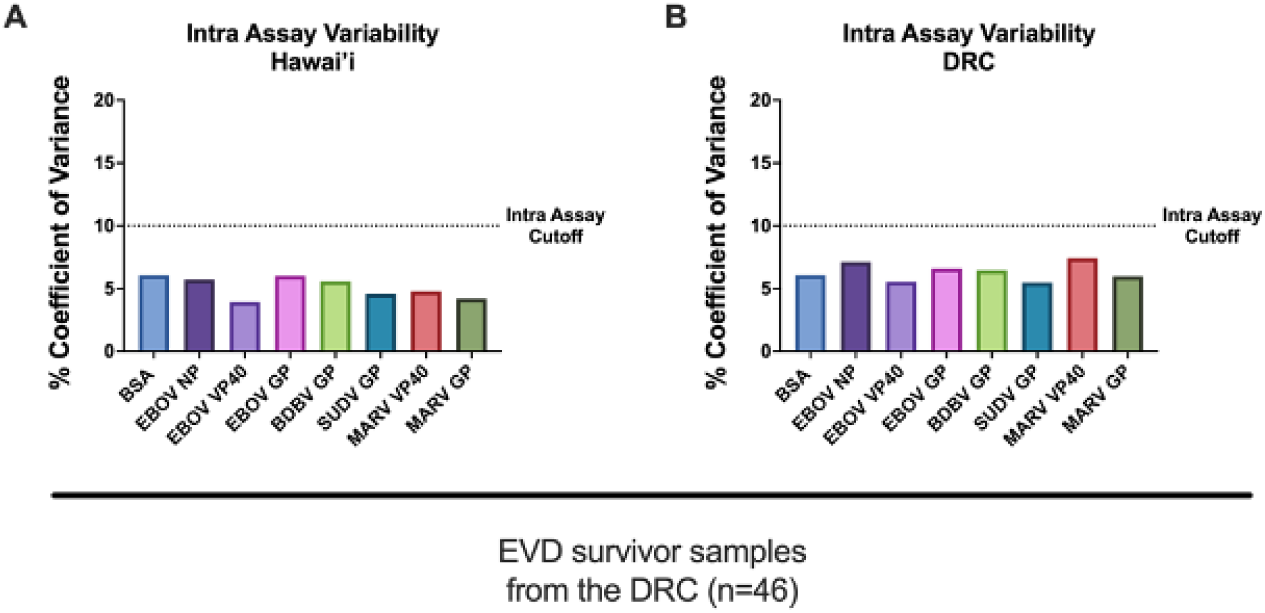
Intra-Assay Variability in Various Geographical and Resourced Settings. (Figure 1 A) Intra-assay variability between duplicates of Yambuku (EVD survivor samples) run at UH. (Figure 1 B) Intra-assay variability between duplicates of Yambuku (EVD survivor samples) run at the INRB. The intra-assay cutoff point is % CV=10.

The EBOV samples were diluted using PBS-BT (1% BSA and 0.02 % Tween 20 in 1x PBS) for IgG assays at 1:100 using 2μL of serum. Approximately 1,250 beads per analyte were incubated with diluted serum samples in black-sided 96-well plates for 3 hours at 37 °C with gentle agitation in the dark. Plates were washed twice with PBS-BT, following incubation with 50μL of 2.5 μg/mL red phycoerythrin (R-PE)-conjugated goat anti-human IgG (Jackson ImmunoResearch Laboratories, West Grove, PA) for 1 hour at 37 °C. After two additional washes, beads were resuspended in MAGPIX drive fluid and analyzed on a MAGPIX Instrument (Luminex, Austin, TX), with a minimum of 50 beads counted per analyte per well. Results were reported as median fluorescence intensity (MFI).

All assay runs were performed by trained personnel following identical protocols at both sites. No formal lot-to-lot bead variability, interference testing, or comparison to alternative serologic platforms (e.g. ELISA) were performed as part of this evaluation study.

#### Quantitative Standard and IgG Concentration Estimation

To support evaluation of analytical range for EBOV GP, an EBOV GP-Binding IgG standard was generated from pooled sera collected from NHPs immunized with a monovalent EBOV GP vaccine. GP-specific IgG standard was purified by immunoaffinity chromatography using EBOV GP-coupled NHS-Sepharose (Cytiva, Marlborough, MA), followed by protein A affinity chromatography. MFI values were plotted against the Log_10_-transformed IgG concentrations and fitted using a sigmoidal dose-response model with variable slope (GraphPad Prism, Boston, MA). Curves meeting r^2^ > 0.99 were used to define the linear portion of the response for estimation of the analytical range.

For prior NHP investigations, GP-specific IgG concentrations for experimental samples were estimated by interpolating sample MFI values against the EBOV GP standard curve and applying dilution factors, with results expressed as µg/mL. Quantitative interpolation was not performed for the human serum samples analyzed in this study; human results are presented as MFI and/or relative to antigen specific reactivity thresholds.

### Analysis

#### Statistical Methods

All statistical analyses were performed using GraphPad Prism version 10 (GraphPad Software, Boston, MA). MFI values were used for all analyses of human serum samples.

For precision and inter-laboratory reproducibility assessments (Cohort 1), coefficients of variation (CV) were calculated for duplicate measurements within laboratories (intra-assay) and between laboratories (inter-laboratory). Pearson correlation coefficients (r^2^) were calculated to assess concordance of analyte specific MFI values generated at the UH and INRB laboratories. For longitudinal analyses of vaccine induced antibody responses (Cohort 2), mean MFI values with standard deviations were calculated at each time point. Comparisons across time points were assessed by one-way or two-way repeated-measures analysis of variance (ANOVA), as appropriate. When pairwise comparisons were conducted, Student’s t-test was performed. All statistical tests were two-sided, and p-values of less than 0.05 were considered statistically significant. Area under the curve (positive incremental) values were calculated using the trapezoidal method relative to the baseline to summarize longitudinal antibody reactivity trends. Statistical analyses were descriptive and comparative in nature and were not intended to assess diagnostic accuracy or clinical sensitivity.

#### Establishment of Antigen-Specific Reactivity Thresholds

Antigen-specific seroreactivity thresholds were established using serum samples from an assumed EBOV-naive population collected in Kinshasa, DRC, in 2017 (n= 379). This assumed-naïve cohort included individuals with no known filovirus exposure history, including participants with no EBOV vaccination, close contact with known cases and/or suspected cases, or prior involvement in an outbreak – all samples were collected in 2017 as part of a larger study as previously described (20–23).

For each antigen target included in the multiplex panel, the reactivity threshold was defined as the mean MFI value with the background BSA value subtracted plus three standard deviations of the assumed-naïve cohort. These thresholds were used to classify samples as reactive or non-reactive for descriptive analyses and visualization of longitudinal antibody responses. Antigen-specific reactivity cut offs with this same MIA have been previously published (24). Reactivity thresholds were not intended to represent diagnostic cutoffs, nor were they validated against an external reference assay. Thresholds were applied uniformly across analyses to facilitate internal comparisons of antibody reactivity within and across cohorts.

## RESULTS

### Assay Precision

To assess intra-assay variability, the variance between duplicate measurements of the 46 Cohort 1 samples was assessed at UH and INRB. Analysis of coefficients of variation (CV) demonstrated comparable assay performance across the two laboratory settings. For all filovirus antigen targets included in the multiplex panel, CV values remained under 10% for duplicate measurements at each site **(Figure 1 A & B)**. Inter-laboratory variability was assessed by comparing analyte-specific results generated at UH and INRB. For all targets, inter-laboratory CV values remained below the predefined threshold of 15% (**Figure 2).** To further evaluate concordance between laboratories, Pearson correlations were calculated for each analyte across sites **(Figure 3)**. Strong linear correlations were observed between sites (r^2^:0.86 and 0.92), with the greatest variance observed in samples approaching the lower limits of detection, supporting the hypothesis of reproducible assay performance across geographic settings.

**Figure 2:**
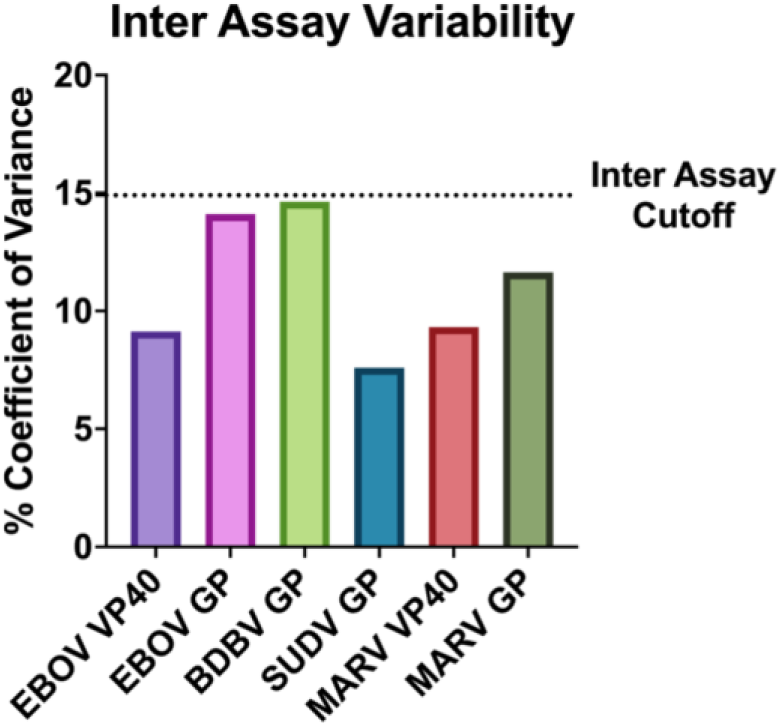
Inter-Assay Variability. Variability presented for each analyte, %CV is recorded between laboratories in Hawaii and DRC. The inter-assay cutoff is 15% CV.

**Figure 3:**
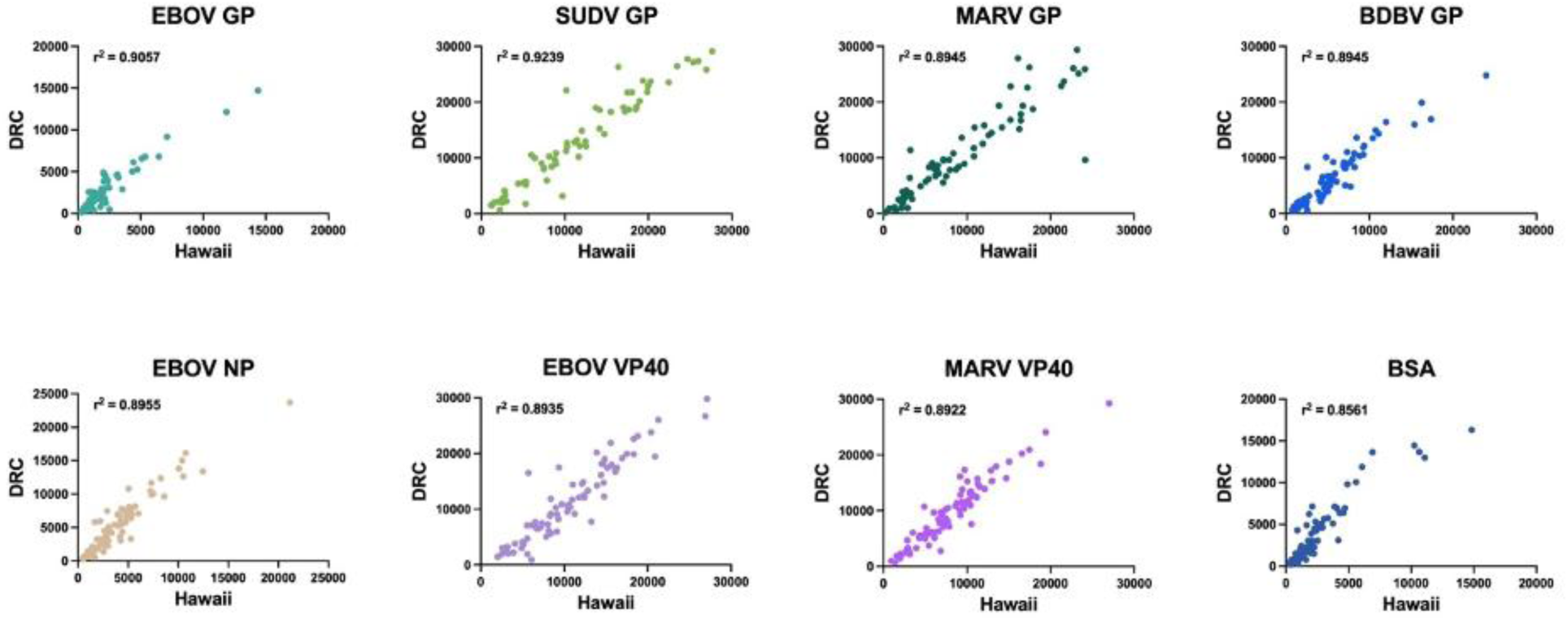
Inter-Assay Variability Correlations by assay location for all filovirus analytes and the control beads (BSA). MFI outputs for each antigen target for every individual sample tested at both UH and INRB laboratory locations (n = 46). Pearson’s correlation is reported for each antigen – higher correlation coefficients highlight the limit intra-assay variability between different sites and operators.

#### Linearity and Range

At the UH site, analytical range was evaluated using the NHP-derived anti-EBOV GP IgG standard. Upper and lower limits of quantification (ULOQ and LLOQ) were defined based on the highest and lowest concentrations within the linear portion of the MFI standard curve. The resulting curve demonstrated a linear range extending from 15.85 ng/mL to 1000 ng/mL (10^1.2^ ng/mL to 10^3^ ng/mL) for NHP samples **(Figure 4)**. These analyses were used to characterize analytical performance for the EBOV GP target only.

**Figure 4:**
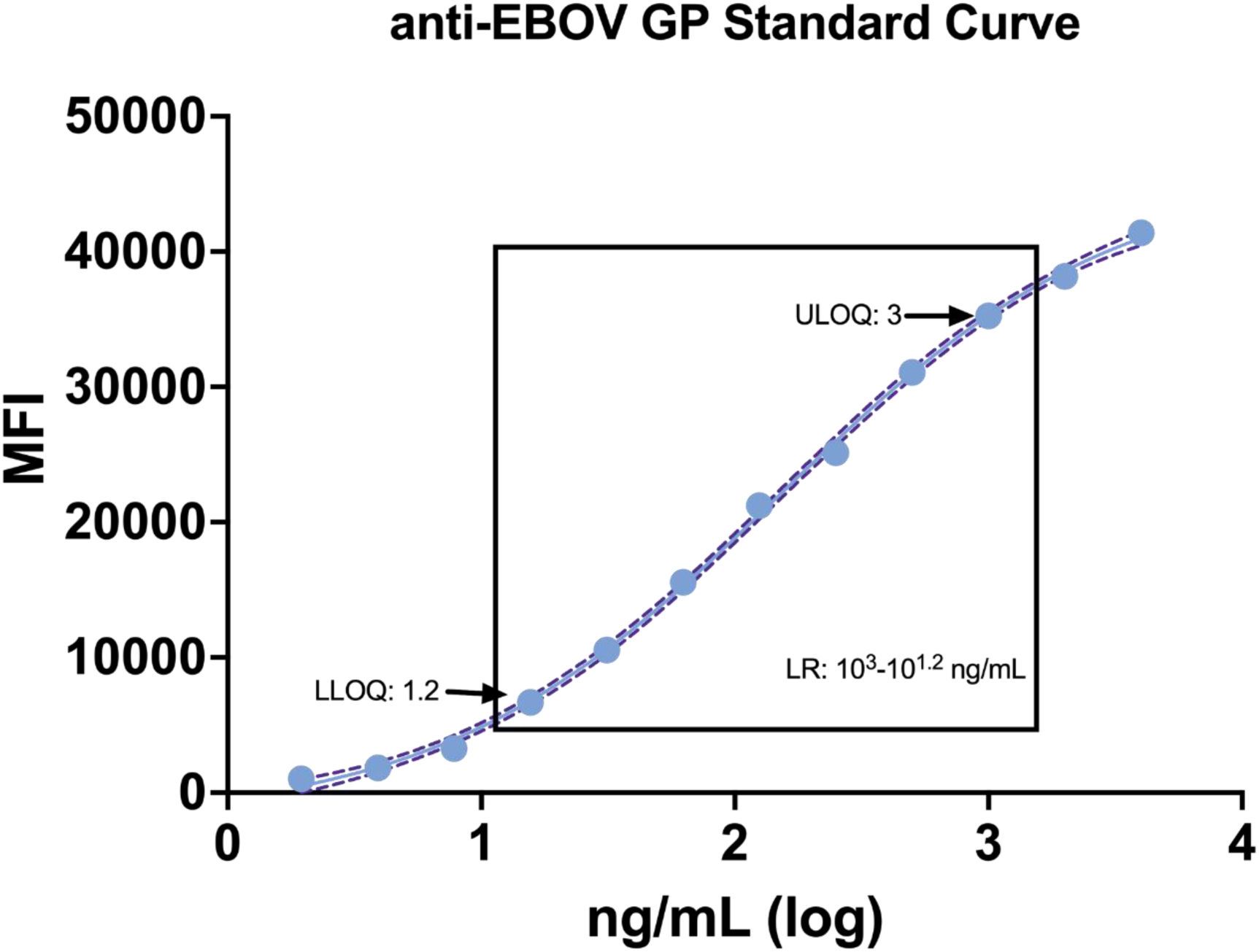
Standard Curve and Linear Range of the NHP EBOV-GP IgG Standard. The standard curve run at UH, derived using NHP EBOV-GP IgG standard, indicates a linear range of 15.85 ng/mL to 1000 ng/mL. The upper (ULOQ) and lower limits (LLOQ) are defined as the upper and lower points of the linear range of the sigmoidal curve for NHP samples allowing quantitation of IgG concentrations.

#### Antigen Discrimination and Assessment of Cross-Reactivity

Following the evaluation of inter-assay and inter-laboratory reproducibility, antibody reactivity patterns were examined among Cohort 2 samples collected longitudinally from individuals in Beni and Mbandaka, DRC, before and after ERVEBO vaccination. Reactivity to EBOV GP and VP40 was assessed up to eight months post immunization. EBOV GP reactivity increased from baseline to 21-days post-vaccination and remained above the antigen-specific reactivity threshold at 8-months in both locations. Conversely, EBOV VP40 reactivity remained stable over time, with mean MFI values consistently below the EBOV VP 40 cutoff point/reactivity threshold. **(Figure 5)**.

**Figure 5:**
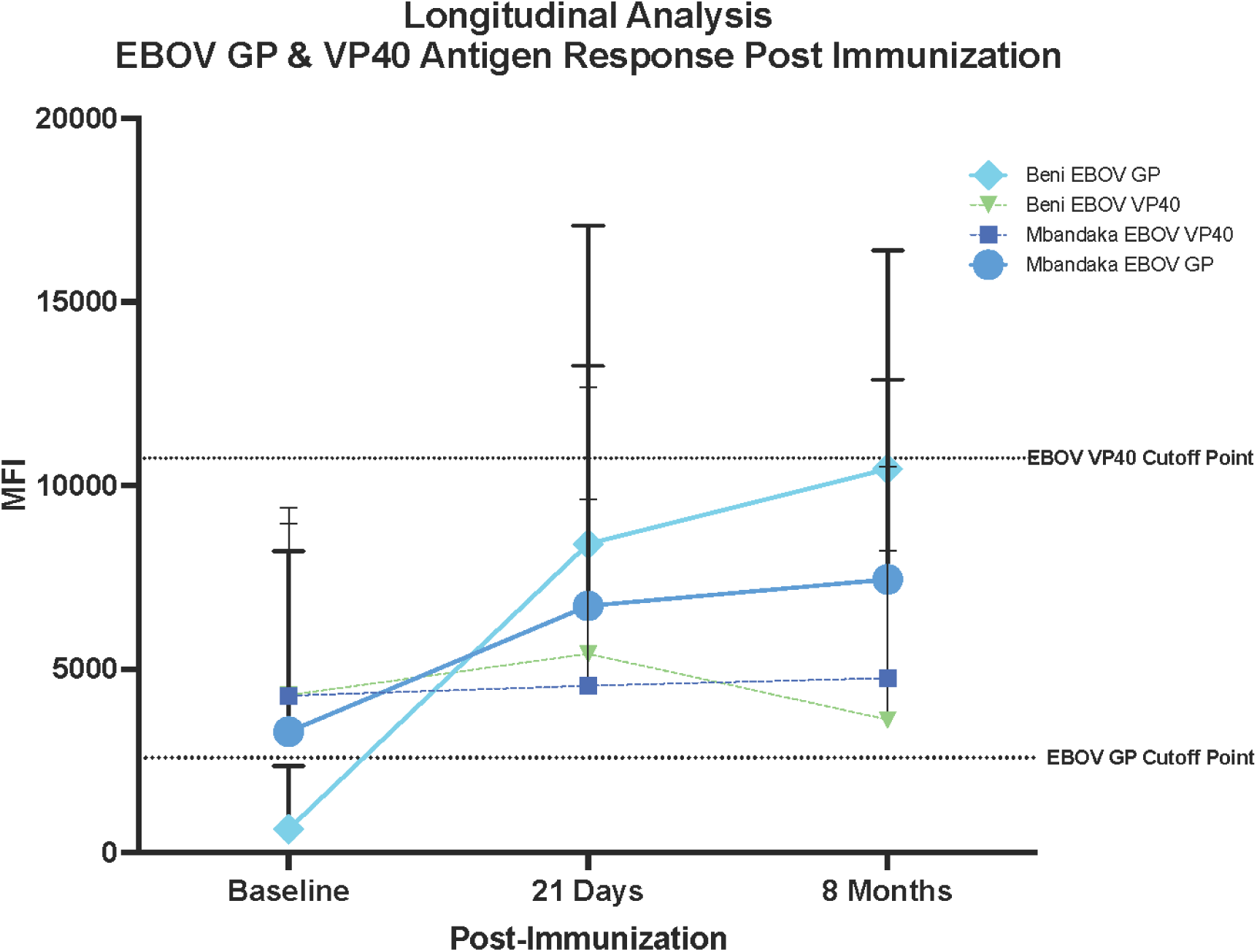
Longitudinal Analysis of EBOV GP & EBOV VP40. EBOV GP reactivity was measured post-immunization at three time points in two health regions of the DRC, Mbandaka, and Beni. Measurements were taken at Baseline (pre-immunization), 21 days, and eight months post-immunization. Data from Mbandaka is shown in dark blues, Beni in green and light blue. Solid lines represent EBOV GP; dashed lines equate to EBOV VP40. Means and SD are shown. EBOV GP cutoff point and EBOV VP40 cutoff point were calculated using Kinshasa naive sample means.

To further examine the antigen discrimination within the multiplex panel, Cohort 2 samples were evaluated for reactivity to MARV, SUDV, and BDBV GP over the same time points. While EBOV GP reactivity increased following vaccination, reactivity to non-EBOV filovirus antigens remained relatively constant and below their respective thresholds across all sampling time points, indicating minimal cross reactivity within this vaccinated cohort (**Figure 6**). BSA-coupled beads were included as an additional target to assess non-specific serum binding and exhibited consistently low MFI values across all collections (mean MFI < 1000), representing a negligible portion of the dynamic range compared to antigen-specific signals, further emphasizing the assay detection of a low background signal.

**Figure 6:**
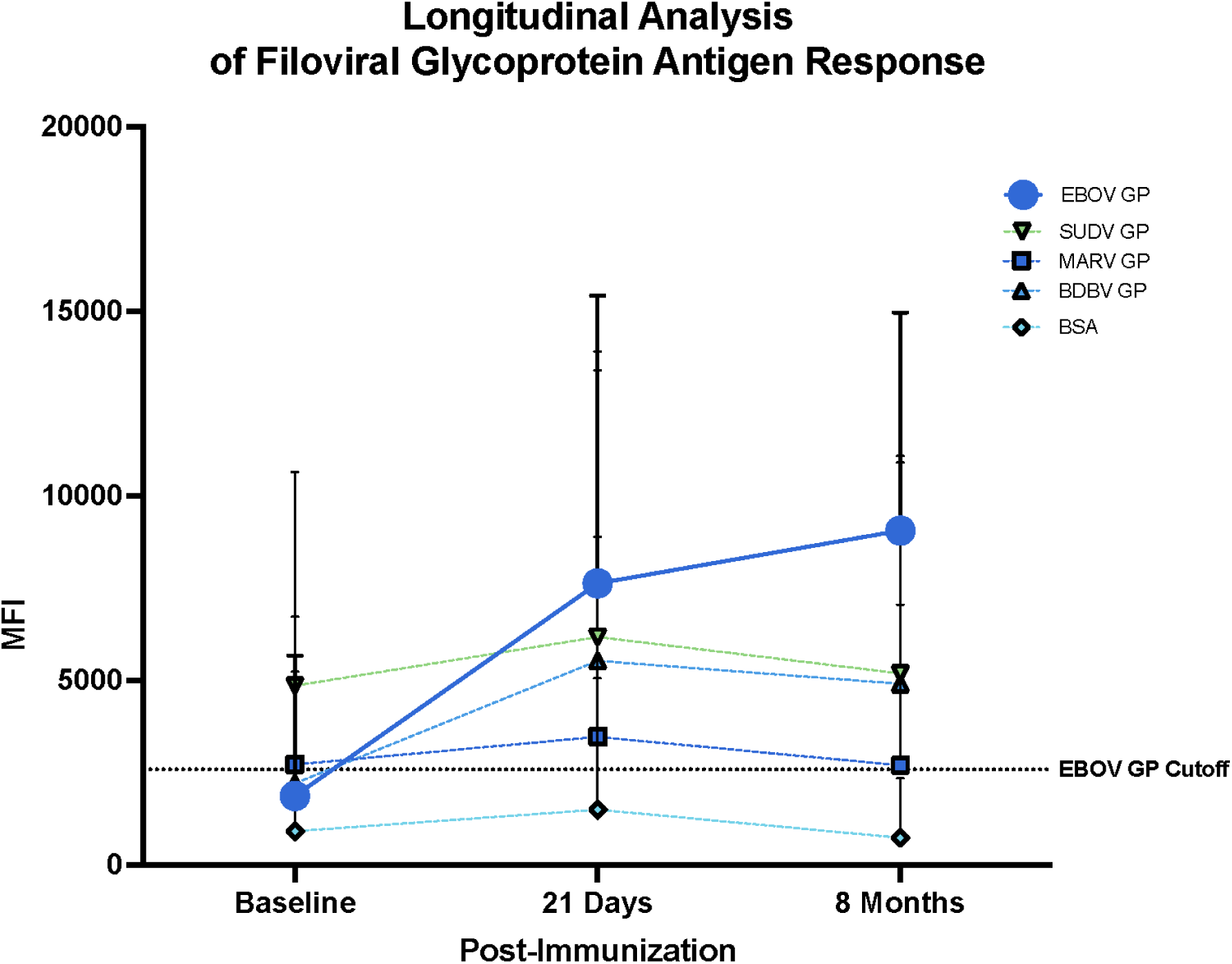
Longitudinal Analysis of Filoviral Antigen Response for Detection of Cross-Reactivity. Pre-immunization (Baseline), 21 days, and eight months post-immunization reactivity to filovirus glycoproteins (EBOV GP), SUDV GP, MARV GP, and BDBV GP from the Mbandaka and Beni cohorts. Negative control, BSA, was used for the baseline assay background. Mean and SD are shown. EBOV GP cutoff point or reactivity threshold—3580 MFI—is indicated by the dashed line.

## DISCUSSION

Through a systematic performance evaluation, we provide important support for the value of MIAs in providing reproducible and consistent results for EBOV antibody detection when implemented across geographically and resource-diverse laboratory settings. Assessment of intra-assay and inter-laboratory variability showed low coefficients of variability and strong concordance between laboratories, supporting the reliability of this assay platform when applied using standardized protocols. In addition, this analysis establishes potential upper and lower limits of the anti-EBOV-GP IgG assay for NHP and possibly human samples, providing reference information relevant to future assay applications and supporting the utility of the assay for serosurveillance and potential immuno-bridging activities. These findings indicate that the MIA can serve as a robust tool for comparative serologic investigations across settings, including low-resource environments, and underscore its potential utility in global health research and response efforts.

Within the context of this evaluation, the multiplex format enabled discrimination between EBOV GP-specific vaccine-induced antibody responses and non-vaccine antigens included in the panel.

Among vaccinated cohorts, increases in EBOV GP reactivity were observed alongside stable reactivity to EBOV VP40 and other filovirus antigens, consistent with expected biological responses to ERVEBO vaccination (25). This pattern supports the utility of the assay for distinguishing antigen-specific responses within a multiplex framework. In addition, the analytical range for EBOV GP was characterized using NHP-derived IgG standard, providing reference information relevant to future assay applications. Quantitative interpolation, however, was not performed for human sera, and further work is required to establish quantitative performance characteristics in human populations.

Assay precision and sustainability are fundamental for validation, particularly for applications involving longitudinal monitoring of vaccine-induced immune responses (26). Assessment of both intra-assay and inter-assay consistency emphasizes the need for performing validation with repeated testing of closely clustered results (27). In this study, reproducible performance was observed across two laboratories operating in distinct geographic and resource contexts. Low intra-assay variability and strong inter-laboratory correlations indicate that assay outputs are stable across sites, despite differences in personnel and operational environments. These findings support the feasibility of deploying standardized multiplex serologic assays across diverse settings for research and surveillance purposes.

A sustainable assay should be able to reliably perform over time, across multiple operators, laboratories, or geographical locations. In evaluating vaccine-induced immune dynamics among the vaccinated Cohort 2, longitudinal changes in anti-EBOV GP antibody seroreactivity were detected over time, while reactivity to EBOV VP40 remained stable. This pattern is consistent with antigen-specific vaccine responses and supports the capacity of the assay to discriminate vaccine induced reactivity within the multiplex panel. The assay also enabled assessment of the durability of the vaccine-induced anti-EBOV GP IgG responses across all three included timepoints. These data illustrate the utility of the MIA for monitoring vaccine-associated immune dynamic but do not constitute an assessment of diagnostic accuracy or clinical effectiveness.

Specificity, in the context of this study, was evaluated through assessments of antigen discrimination and lack of cross-reactivity within vaccinated cohorts. Comparison of relative reactivity between EBOV GP and VP40 following vaccination demonstrated expected antigen-specific response patterns. Additional evaluation of reactivity to other filovirus GP targets (MARV, SUDV, BDBV) showed that mean MFI values for non-EBOV GP antigens remained below their respective reactivity thresholds. These findings indicate minimal cross-reactivity within the vaccinated population and are consistent with the expected antigenic specificity of the ERVEBO vaccine. However, specificity for non-EBOV targets was inferred from the absence of increased reactivity rather than confirmed using known positive sera.

Several limitations should be considered when interpreting these findings. Logistically, while MIA is robust, its implementation in LMICs is still tethered to specialized instrumentation and the need for a stable power supply and trained personnel. Analytically, our quantitative performance evaluation relied on an NHP-derived EBOV IgG standard, without external platform comparison (e.g. ELISA). While the performance of the EBOV GP target alone has been extensively compared to the gold standard Filovirus Animal Non-Clinical Group (FANG) assay, biological validation was restricted by the unavailability of confirmed non-EBOV positive human sera (28).

Consequently, specificity for these targets was inferred from the lack of cross-reactivity from individuals with other common regional infections or non-EBOV filovirus infections, rather than confirmed through known exposure history and detected reactivity.

This performance evaluation provides evidence that a multiplex bead-based immunoassay can be reproducibly implemented across diverse laboratory settings and can support antigen-specific serologic analyses within a filovirus-focused panel. High-impact and economical assays, such as the MIA, can have an influence on the crafting of public health interventions, such as real-time monitoring of vaccine coverage, or identifying transmission in areas where clinical surveillance is sparse, and policies, particularly in resource-limited settings where disease surveillance and vaccine coverage estimates may be largely unknown in many contexts. Evaluations using a validation methodology, including expanded assessment of non-EBOV targets and quantitative performance in human populations, will further strengthen the utility of this platform for global health research, serosurveillance, and vaccine response monitoring.

## Disclaimer

The views and conclusions contained in this document are those of the authors and should not be interpreted as necessarily representing the official policies, either expressed or implied, of the U.S. Department of Health and Human Services or of the institutions and companies affiliated with the authors, nor does mention of trade names, commercial products, or organizations imply endorsement by the U.S. Government. This project has been funded in whole or in part with Federal funds from the Food and Drug Administration (Grant No. 75F40119C10128) and the Gates Foundation (OPP1195609).

## Data Availability

All data produced in the present study can be made available upon reasonable request to the corresponding author.

